# Myositis associated anti-NT5C1A autoantibody in clinical practice

**DOI:** 10.1101/2020.03.25.20043760

**Authors:** Chiseko Ikenaga, Andrew R. Findlay, Namita A. Goyal, Sarah Robinson, Jonathan Cauchi, Yessar Hussain, Leo H. Wang, Joshua C. Kershen, Brent A. Beson, Michael Wallendorf, Robert C. Bucelli, Tahseen Mozaffar, Alan Pestronk, Conrad C. Weihl

**Affiliations:** Department of Neurology, Washington University School of Medicine, St. Louis, MO; Department of Neurology, University of California, Irvine, CA; Austin Neuromuscular Center, The University of Texas Dell Medical School, Austin, TX; Department of Neurology, University of Washington, Seattle, WA; Integris Southwest Medical Center, Oklahoma City, OK; Division of Biostatistics, Washington University School of Medicine, St. Louis, MO

**Keywords:** anti-NT5C1A autoantibody, inclusion body myositis, dermatomyositis, immune-mediated necrotizing myopathy, antisynthetase syndrome

## Abstract

**Objective:** To define the diagnostic utility and clinicopathologic features correlating with anti-cytosolic 5′-nucleotidase 1A (NT5C1A) antibody positivity in idiopathic inflammatory myopathies (IIMs).

**Methods:** 4987 patients had anti-NT5C1A status clinically tested between 2014 and 2019 in the Washington University neuromuscular clinical laboratory. Using clinicopathologic information available for 630 of these patients, we classified them as inclusion body myositis (IBM), dermatomyositis, antisynthetase syndrome, immune-mediated necrotizing myopathy (IMNM), non-specific myositis, or non-inflammatory muscle diseases.

**Results:** Anti-NT5C1A was found in 182/287 patients with IBM (63%), 11/53 patients with dermatomyositis (21%), 7/27 patients with antisynthetase syndrome (26%), 9/76 patients with IMNM (12%), 20/84 patients with non-specific myositis (24%), and 6/103 patients with non-inflammatory muscle diseases (6%). The sensitivity and specificity of anti-NT5C1A seropositivity for the diagnosis of IBM was 63% and 85%, respectively. Anti-NT5C1A positive IBM patients had a higher frequency of finger flexion weakness (*p*<0.01) and their biopsies harbored more COX negative fibers (*p*=0.04). 18% of anti-NT5C1A negative IBM patients seroconverted to anti-NT5C1A positive on repeat testing.

**Conclusions:** This is the largest description of patients tested by a clinical diagnostic lab for anti-NT5C1A. We confirm the sensitivity and specificity of anti-NT5C1A for IBM and identified clinicopathologic features in IBM which correlate with anti-NT5C1A status. Notably, anti-NT5C1A testing increased both the diagnostic sensitivity and specificity of IBM when combined with patient age, gender and creatine kinase (CK) level. We propose that future IBM diagnostic criteria include anti-NT5C1A testing.

## INTRODUCTION

Inclusion body myositis (IBM) is an idiopathic inflammatory myopathy (IIM) that typically affects patients over the age of 50 [1]. Patients with IBM are clinically characterized by asymmetric finger flexion and knee extension weakness [1]. In 2013, anti-cytosolic 5′-nucleotidase 1A (NT5C1A) antibody was detected in the sera of patients with IBM and recognized as a potential diagnostic marker for IBM [2, 3]. Subsequently, the antibody was detected in patients with dermatomyositis, Sjögren’s syndrome, and systemic lupus erythematosus [4, 5, 6, 7, 8, 9, 10]. This suggests that anti-NT5C1A can be detected in autoimmune diseases other than IBM. The relationship between seropositivity for anti-NT5C1A and other clinicopathologic features in IBM or dermatomyositis have been discussed and some report that seropositivity for anti-NT5C1A in IBM or juvenile myositis predict a more severe phenotype [4, 10, 11, 12]. However, this relationship has not been assessed in other IIMs.

Diagnostic testing for myositis specific and myositis associated antibodies are routinely performed in patients with suspected IIM. However, their sensitivity, specificity and clinical utility vary from testing lab to testing lab. This feature makes it essential to describe testing lab specific data and its relation to clinical practice. Since 2014, the Washington University neuromuscular clinical laboratory has tested anti-NT5C1A via Western blot on 4987 patients, from >100 distinct clinical institutions, who were suspected of neuromuscular disease. 1401 patients (28%) were seropositive for anti-NT5C1A. In this study, we aimed to confirm anti-NT5C1A’s utility in diagnosing IBM, clarify the relationship between seropositivity for anti-NT5C1A and other clinicopathologic features in IIMs, and determine the usefulness of the antibody in clinical practice.

## METHODS

### Patients

We retrospectively identified 4987 patients, who underwent anti-NT5C1A testing at Washington University in St. Louis from 2014 to 2019.

The clinical chart, biopsy reports, and results of autoantibodies status of 1210 patients were then reviewed from Washington University School of Medicine in St. Louis, University of California, Irvine, The University of Texas Dell Medical School, University of Washington, and Integris Southwest Medical Center (Fig.1). We excluded those patients with diseases whose primary pathology is outside of skeletal muscle or whose final diagnosis was still under investigation.

**Figure1:**
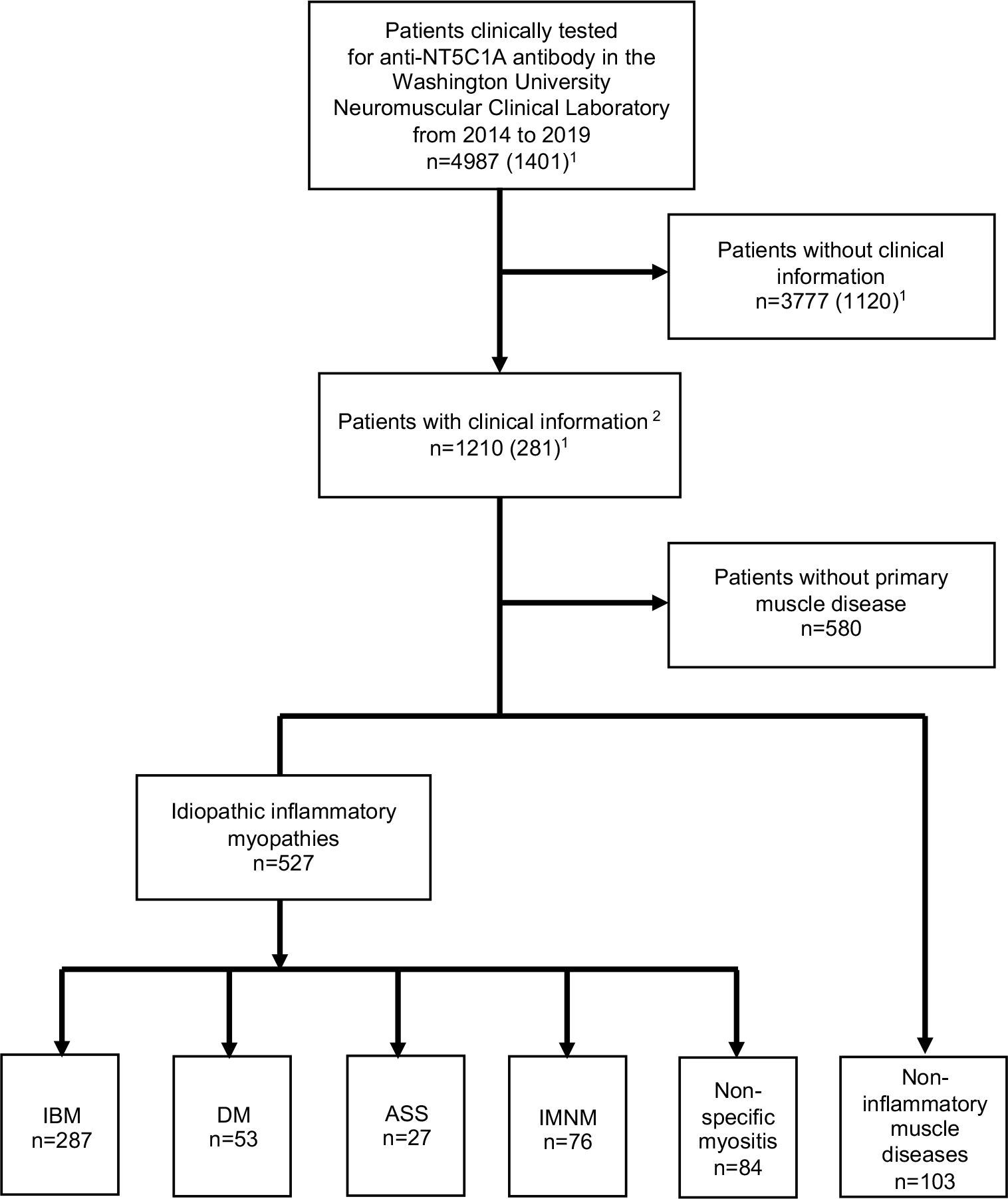
Flowchart of patients in this study. There were 1210 patients with clinical information from five institutes. For the diagnosis of IBM, we used the diagnostic criteria and reached consensus of the European Neuromuscular Center (ENMC). We also included IBM patients, who had clinical features, yet no muscle biopsy was performed. For the diagnosis of dermatomyositis and IMNM, we also used the diagnostic criteria and reached consensus of the ENMC. We classified those with anti-aminoacyl tRNA synthetase antibody in addition to one or more of the following clinical features (Raynaud’s phenomenon, arthritis, interstitial lung disease, fever, mechanic’s hands or biopsy finding of immune myopathies with perimysial pathology) as anti-synthetase syndrome. ^1^: The numbers in the parentheses indicates the number of patients, who were seropositive for anti-NT5C1A. ^2^: Washington University School of Medicine in St. Louis, University of California, Irvine, The University of Texas Dell Medical School, University of Washington, Integris Southwest Medical Center. IBM=inclusion body myositis; DM=Dermatomyositis; ASS=Antisynthetase syndrome; IMNM=immune-mediated necrotizing myopathy.

IBM was classified by European Neuromuscular Center (ENMC) criteria [13]. 37 patients with definite or probable clinical criteria of IBM; yet a muscle biopsy had not been performed were also included. ENMC diagnostic criteria was used for the diagnosis of dermatomyositis and immune-mediated necrotizing myopathy (IMNM) [14,15]. Anti-synthetase syndrome was classified by the following, the presence of anti-aminoacyl tRNA synthetase antibody and one or more of the following clinical features (Raynaud’s phenomenon, arthritis, interstitial lung disease, fever (not attributable to another cause), mechanic’s hands or biopsy finding of immune myopathies with perimysial pathology) [16, 17, 18]. In this way, the cohort included patients with IBM (n=287), dermatomyositis (n=53), antisynthetase syndrome (n=27), and IMNM (n=76). Patients that did not meet this criteria and had a muscle biopsy with endomysial or perimysial inflammation, MHC-class 1 up-regulation on muscle fibers or complement C5b-9 complex deposits on endomysial capillaries were categorized as non-specific myositis (n=84).

Patients with a non-inflammatory, hereditary or presumed hereditary muscle disorder were also included. The diagnoses of these patients were VCP-related multi-system proteinopathy, limb-girdle muscular dystrophy, Becker muscular dystrophy, myotonic dystrophy, centronuclear myopathy, myofibrillar myopathy, mitochondrial myopathy, rhabdomyolysis, and McArdle’s disease (n=103).

### Serological testing for autoantibodies

Anti-NT5C1A testing was clinically performed by the Washington University Neuromuscular Laboratory via Western blotting as previously described [11]. In some cases, anti-Jo-1 antibody, anti-HMGCR antibody, anti-SRP antibody, and anti-MDA5 antibody were also performed by Washington University Neuromuscular Laboratory [19]. Serums were tested and interpreted blindly without knowledge of the diagnosis. All other antibody data was extracted from the clinical chart. Panels of serum muscle specific antibodies were evaluated by clinical laboratories in Oklahoma or California.

### Myopathologic analysis

The percentages of cytochrome oxidase (COX) negative and succinate dehydrogenase (SDH) positive muscle fibers were determined by photographing a random field with a 10x objective and counting 200 fibers. A cutoff value of 1.0% was used for qualitative analysis of COX negative fibers as with our previous report [20].

### Definition of clinical features and assessment of responses to immunotherapy

Dermatologic features were defined by clinical description (i.e., Heliotrope rash, Gottron’s sign, V-sign, shawl sign, calcinosis cutis, and mechanic’s hands), skin biopsy data, and/or expert opinion of a dermatologist. Inflammatory arthritis was defined by imaging features and/or expert opinion of a rheumatologist. Interstitial lung disease was defined by imaging features, pulmonary function tests, biopsy data, and/or expert opinion of a pulmonologist.

Responses to treatments were determined for patients followed by more than 1 year as: effective, muscle weakness had diminished and the level of activities of daily living had improved to the same as premorbid level; no worsening, some symptoms had improved but there were still several symptoms or abnormality on the blood test; worsening, muscle weakness was progressive and there was no improvement with any immunosuppressive treatments.

### Statistical analyses

Fisher exact test compared categorical variables and t-test compared continuous variables. We considered *p* values less than 0.05 as statistically significant. To control for multiple comparisons, we applied Bonferroni correction to provide an adjusted threshold for significance in Table 3 and Table 4. Multiple logistic regression models were used to determine the diagnostic utility of anti-NT5C1A for IBM versus IIMs.

**Table 1.**
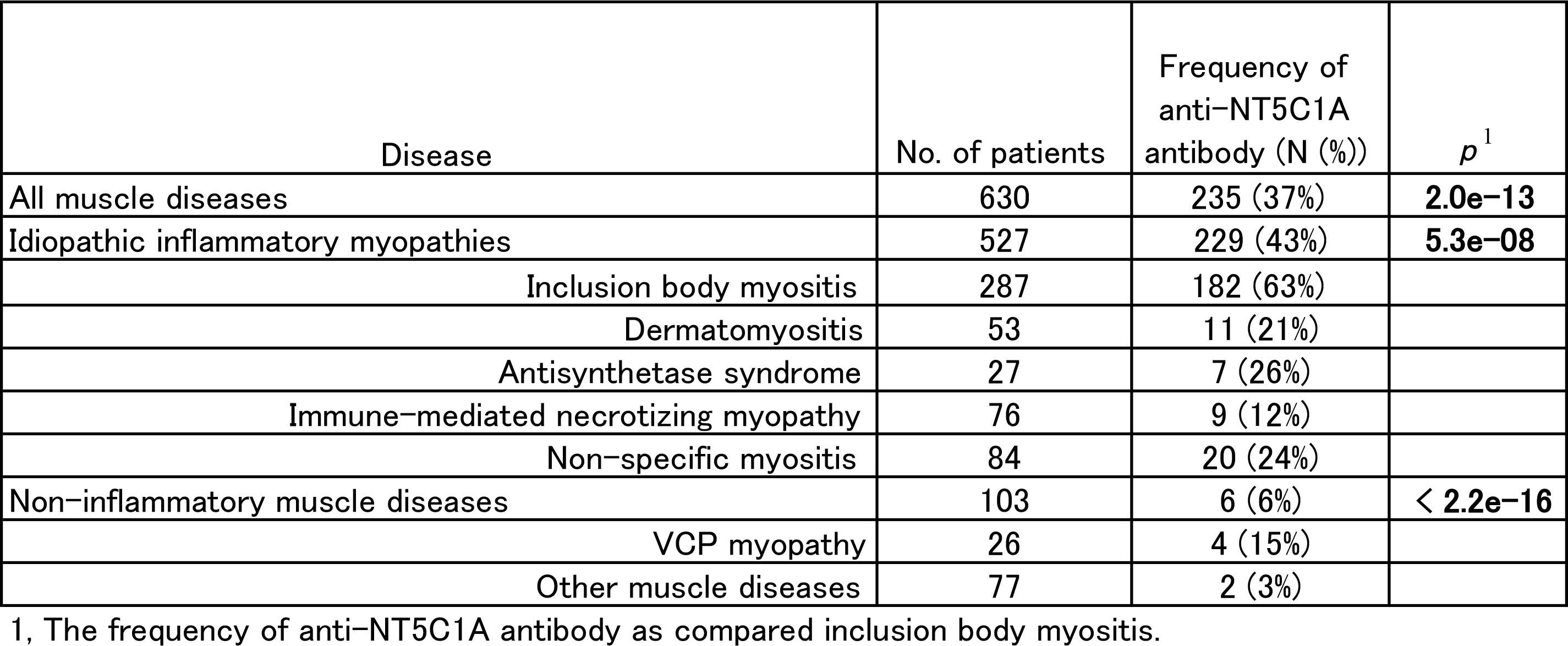
Frequency of anti-NT5C1A antibody in muscle disease

| Disease | No. of patients | Frequency of anti-NT5C1A antibody (N (%)) | $p^1$ |
| --- | --- | --- | --- |
| All muscle diseases | 630 | 235 (37%) | **2.0e-13** |
| Idiopathic inflammatory myopathies | 527 | 229 (43%) | **5.3e-08** |
| Inclusion body myositis | 287 | 182 (63%) |  |
| Dermatomyositis | 53 | 11 (21%) |  |
| Antisynthetase syndrome | 27 | 7 (26%) |  |
| Immune-mediated necrotizing myopathy | 76 | 9 (12%) |  |
| Non-specific myositis | 84 | 20 (24%) |  |
| Non-inflammatory muscle diseases | 103 | 6 (6%) | **< 2.2e-16** |
| VCP myopathy | 26 | 4 (15%) |  |
| Other muscle diseases | 77 | 2 (3%) |  |

1, The frequency of anti-NT5C1A antibody as compared inclusion body myositis.

**Table 2.** Clinicopathologic features of patients with idiopathic inflammatory myopathies

1, The frequency of anti-NT5C1A antibody as compared inclusion body myositis. Table 2, Clinicopathologic features of patients with idiopathic inflammatory myopathies
|  | Patients with idiopathic inflammatory myopathies<br>(n=527) |  |  | Patients with idiopathic inflammatory myopathies<br>(n=527) |  |  | Patients with inflammatory myopathies other than IBM<br>(n=240) |  |  |
| --- | --- | --- | --- | --- | --- | --- | --- | --- | --- |
|  | Anti-NT5C1A positive | Anti-NT5C1A negative | p | IBM | non-IBM | p | Anti-NT5C1A positive | Anti-NT5C1A negative | p |
| Number of patients (M:F) | 229 (120 : 109) | 298 (150 : 148) | 0.66 | 287 (172 : 115) | 240 (98 : 142) | 0 | 47 (13 : 34) | 193 (85 : 108) | 0.05 |
| Age at onset, mean ± SD years | 58.2±10.4 | 56.0±14.8 | 0.05 | 59.9±9.2 | 53.5±15.9 | 0.00000008 | 54.3±14.0 | 53.3±16.4 | 0.67 |
| Maximum creatine kinase level (IU/L) | 895±1508 | 2517±6016 | 0.00002 | 581±632 | 3271±6635 | 0.000000007 | 1912±2809 | 3607±7244 | 0.01 |
| Dysphagia | 100/207 (48%) | 84/222 (38%) | 0.03 | 139/279 (50%) | 45/150 (30%) | 0.0001 | 7/41 (17%) | 31/170 (18%) | 1 |
| Antinuclear antibody | 37/62 (60%) | 49/99 (49%) | 0.26 | 35/74 (47%) | 51/87 (59%) | 0.16 | 14/31 (45%) | 22/121 (18%) | 0.0036 |
| Other antibodies<sup>1</sup> | 47/127 (37%) | 127/180 (71%) | 0.000000007 | 46/153 (30%) | 128/154 (83%) | 0 | 22/47 (47%) | 96/193 (50%) | 0.75 |
| Muscle biopsy findings |  |  |  |  |  |  |  |  |  |
| Endomysial inflammation | 161/177 (91%) | 112/188 (60%) | 0 | 228/246 (93%) | 45/119 (38%) | 0 | 23/38 (61%) | 54/160 (34%) | 0.003 |
| Perimysial inflammation | 54/137 (39%) | 54/170 (32%) | 0.19 | 64/190 (34%) | 44/117 (38%) | 0.54 | 15/38 (39%) | 66/158 (42%) | 0.86 |
| MHC-class1 upregulation | 91/103 (88%) | 109/146 (75%) | 0.0092 | 129/148 (87%) | 71/101 (70%) | 0.0018 | 24/32 (75%) | 88/138 (64%) | 0.3 |

Abbreviations: IBM = inclusion body myositis; MHC = major histocompatibility complex. 1: Antibodies against MDA5, TIF1 γ , NXP2, Mi2, Jo-1, EJ, OJ, HMGCR, SRP, SS-A, SS-B, PM-Scl, and U1snRNP were included.

**Table 3.** Clinicopathologic features of 287 patients with IBM according to anti-NT5C1A antibody status

|  | Anti-NT5C1A positive | Anti-NT5C1A negative | *p*<sup>1</sup> |
| --- | --- | --- | --- |
| Number of patients (M:F) | 182 (107:75) | 105 (65:40) | 0.62 |
| Classification by the ENMC IBM diagnostic criteria 2011 |  |  |  |
| Clinico-pathologically defined IBM (n) | 44 | 44 |  |
| Clinically defined IBM (n) | 85 | 31 |  |
| Probable IBM (n) | 29 | 17 |  |
| Clinically diagnosed IBM (without biopsy, n) | 24 | 13 |  |
| Age at onset, mean ± SD years | 59.2±9.0 | 61.1±9.5 | 0.1 |
| Onset to biopsy, mean ± SD years | 5.2±4.6 | 5.7±5.2 | 0.5 |
| Maximum creatine kinase level (IU/L) | 620±664 | 516±570 | 0.18 |
| Dysphagia | 91/181 (50%) | 48/98 (49%) | 0.9 |
| Knee extension weakness (≥ hip flexion weakness) | 137/182 (75%) | 76/105 (72%) | 0.68 |
| Finger flexion weakness ($>$ shoulder abduction weakness) | 168/182 (92%) | 81/105 (77%) | **0.0005** |
| Serum autoantibodies other than anti-NT5C1A antibody |  |  |  |
| antinuclear antibody | 24/43 (56%) | 11/31 (35%) | 0.1 |
| anti-SSA (Ro) antibody | 10/47 (21%) | 6/28 (21%) | 1 |
| anti-SSB (La) antibody | 3/47 (6%) | 1/28 (4%) | 1 |
| anti-PM-Scl antibody | 1/47 (2%) | 2/28 (7%) | 0.55 |
| anti-U1snRNP antibody | 1/47 (2%) | 1/28 (4%) | 1 |
| anti-Jo-1 antibody | 5/75 (7%) | 6/48 (13%) | 0.33 |
| anti-HMGCR antibody | 10/78 (13%) | 10/51 (20%) | 0.33 |
| anti-SRP antibody | 3/45 (7%) | 0/31 | 0.27 |
| anti-MDA5 antibody | 2/43 (5%) | 3/25 (12%) | 0.35 |
| Muscle biopsy findings |  |  |  |
| Vacuoles | 93/153 (61%) | 65/91 (71%) | 0.1 |
| Protein accumulation | 71/95 (75%) | 55/66 (83%) | 0.24 |
| Endomysial inflammatory infiltrate | 147/155 (95%) | 81/91 (89%) | 0.13 |
| Focal invasion of non-necrotic fibers | 84/117 (72%) | 44/72 (61%) | 0.15 |
| MHC-class1 upregulation | 78/85 (92%) | 51/63 (81%) | 0.08 |
| Qualitative analysis of COX negative fibers<sup>2</sup> | 86/122 (70%) | 53/77 (69%) | 0.87 |
| COX negative fibers<sup>2</sup> (%) | 3.2±3.9 | 2.1±2.3 | **0.04** |
| SDH positive fibers (%) | 2.2±2.9 | 2.0±2.8 | 0.69 |
| Complications of other autoimmune disease | 24/180 (13%)<sup>3</sup> | 18/97 (19%) | 0.29 |
| Sjögren syndrome | 7 | 4 |  |
| Rheumatoid arthritis | 2 | 4 |  |
| Systemic lupus erythematosus | 3 | 0 |  |
| Scleroderma | 2 | 1 |  |
| Hypothyroidism | 15 | 9 |  |
| Sarcoidosis | 1 | 4 |  |
| Malignancy<sup>4</sup> | 15<sup>5</sup> | 8<sup>6</sup> |  |

Abbreviations: ENMC = European Neuromuscular Center; COX = cytochrome oxidase; SDH = succinate dehydrogenase.
1: Bonferroni corrected significance thresholds for clinical features and biopsy findings were 0.003 and 0.007, respectively.
2: The percentages of COX negative and SDH positive muscle fibers were determined by photographing a random field with a 10x objective and counting 200 fibers. We set cutoff value as 1.0% for qualitative analysis of COX negative fibers.
3: Five patients had two autoimmune diseases (One patient had Sjögren syndrome and systemic lupus erythematosus, one patient had rheumatoid arthritis and systemic lupus erythematosus, one patient had rheumatoid arthritis and hypothyroidism, two patients had Sjögren syndrome and hypothyroidism.)
4: Not all the patients were tested for CT, PET, or tumor markers as cancer screening.
5: Three breast cancer, two prostate cancer, thyroid cancer, parathyroid adenoma, hepatocellular cancer, Paget's disease, non-Hodgkin's lymphoma, lymphoplasmacytic lymphoma, Kaposi's sarcoma, basal cell carcinoma of the forehead, small cell carcinoma of right main bronchus, squamous cell carcinoma of unknown primary.
6: Two T-cell large granular lymphocyte leukemia, two prostate cancer, meningioma, Hodgkin's lymphoma, basal cell carcinoma in legs, basal cell carcinoma of skin.

**Table 4.**
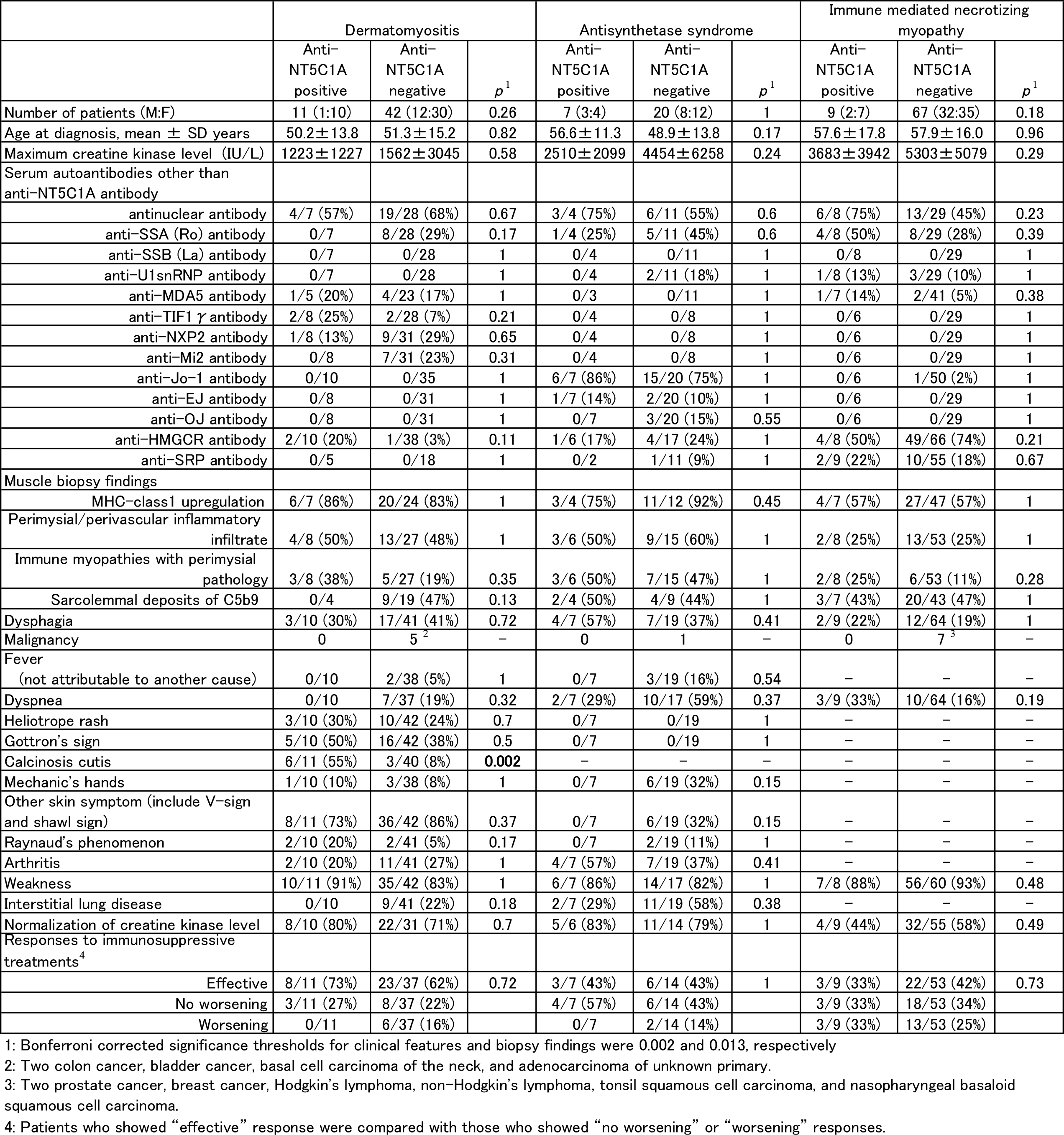
Clinicopathologic features of patients with dermatomyositis, antisynthetase syndrome, and immune mediated necrotizing myopathy according to anti-NT5C1A status

### Protocol approvals and patient consents

All studies were approved by the Human Studies Committee at Washington University in St. Louis.

### Data availability

The data that support the findings of this study are available from the corresponding author, upon reasonable request.

## RESULTS

Among 527 patients with clinical or pathologically defined myositis, 229 patients (43%) were seropositive for anti-NT5C1A of which 182 patients (79%) were classified as IBM (Table1). Among all 287 IBM patients, 63% were seropositive for anti-NT5C1A. The sensitivity of anti-NT5C1A seropositivity for the diagnosis of IBM was 63%. Anti-NT5C1A seropositivity was 85 % specific for IBM among all patients with muscle diseases, and 80% specific for IBM among IIMs (Table 1). To understand how anti-NT5C1A testing changes existing sensitivity and specificity using minimal clinical criteria (age, gender and creatine kinase (CK) value) within our 527 patient IIM cohort, we performed multiple logistic regression model analysis (Fig. 2). The addition of anti-NT5C1A status increased the AUC from 0.76 to 0.84 demonstrating an improvement in diagnostic accuracy (Fig. 2 compared curve 1 (age, gender, CK) to curve 2 (anti-NT5C1A, age, gender and CK)). Logistic regression model with the addition of a biopsy with or without endomysial inflammation further increased the AUC to 0.92 (Fig. 2 curve 3 (age, gender, CK, anti-NT5C1A status, and +/- endomysial inflammation)).

**Figure 2:**
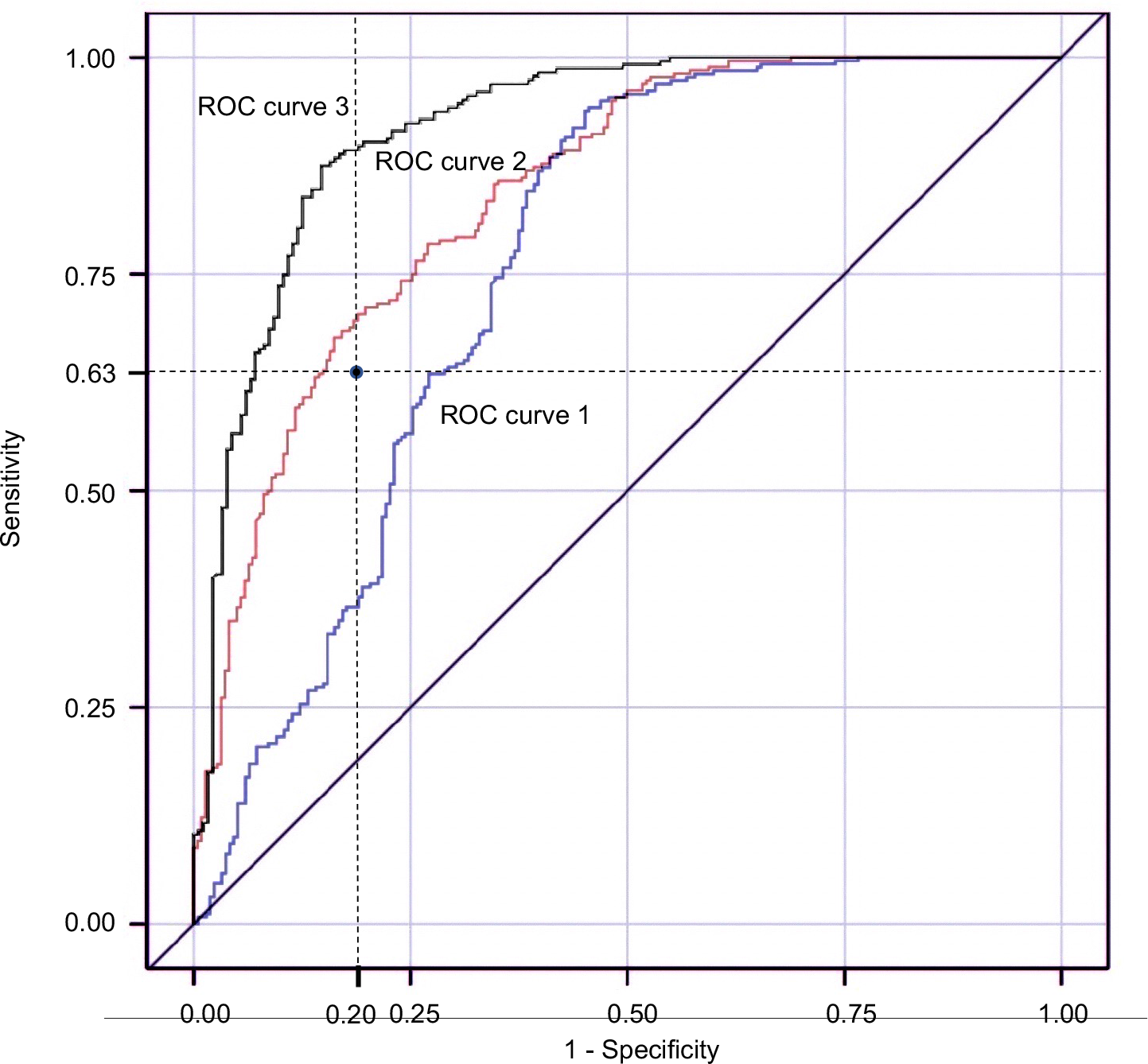
The change of receiver operator characteristic (ROC) curve depending on the information of anti-NT5C1A status. Among the 527 patients with idiopathic inflammatory myopathies, we assessed the utility of anti-NT5C1A antibody for the diagnosis of IBM. The single data point shows the sensitivity of anti-NT5C1A seropositivity for the diagnosis of IBM (63%) while its specificity among patients with IIMs was 80%. ROC curve 1 was based on the following information: age at onset (OR (odds ratio) 1.035, 95% CI 1.018-1.053); male gender (OR 2.398, 95% CI 1.580-3.639); maximum creatine kinase level (OR 0.999, 95% CI 0.999-1.000). ROC curve 2 was based on the information of anti-NT5C1A status (OR 6.473, 95% CI 4.026-10.408), in addition to age at onset, gender, and maximum creatine kinase level. The AUC was larger in ROC curve 2 (0.84) compared with that in ROC curve 1 (0.76). When the limited biopsy finding of endomysial inflammation was included, the model was further improved to the ROC curve 3, whose AUC was 0.92. ROC= receiver operator characteristic; AUC= area under the curve

To understand the differences between anti-NT5C1A seropositive and seronegative patients independent of IIM subtype, we compared these two groups (Table 2). There was no significant difference as to the ratio of males to females, age at onset or antinuclear antibody seropositivity. In contrast, anti-NT5C1A positive patients had a lower serum CK (868±1339 IU/L), higher frequency of endomysial inflammation (91%), and presence of MHC-class1 expression (88%), compared with seronegative patients (*p*<0.01). There was no difference in the presence of perimysial inflammation between the two groups. Notably, complaints of dysphagia were significantly enriched in anti-NT5C1A patients (*p*=0.03).

To see if these differences were due to the overrepresentation of IBM patients in the anti-NT5C1A positive group, we performed a similar analysis that removed the 287 patients with IBM and thus looked at only non-IBM IIMs. Similar to the previous group, anti-NT5C1A positive non-IBM IIM patients had a lower serum CK (1912±2809 IU/L; *p*=0.01) and a higher frequency of endomysial inflammation (61%) suggesting that these two features are not affected by the increased number of IBM patients in the anti-NT5C1A positive group. As expected, independent of anti-NT5C1A status, patients with IBM were significantly older (*p*<0.01) and male 60% as compared to 41% in the non-IBM cohort (*p*<0.01). In addition, IBM patients showed lower serum CK (581±632 IU/L), higher frequency of endomysial inflammation (93%), MHC-class1 expression (87%) and dysphagia (50%).

Anti-NT5C1A positive IBM patients had a higher frequency of finger flexion weakness (*p*<0.01), but no significant differences were seen with regard to age at onset, maximum creatine kinase level, dysphagia, complications of other autoimmune diseases/sarcoidosis/malignancy, or co-existing autoantibodies (Table 3). Anti-NT5C1A positive IBM patients also had significantly more COX negative fibers (*p*=0.04). There were no other significant differences in muscle biopsy features.

We also compared clinicopathologic features and responsiveness to immunosuppressive treatments in non-IBM IIM patients who underwent anti-NT5C1A testing. Among patients with dermatomyositis (n=53), antisynthetase syndrome (n=27), and IMNM (n=76), 11 (21%), 7 (26%), and 9 (12%) patients were seropositive for anti-NT5C1A, respectively (Table 4). Dermatomyositis patients seropositive for anti-NT5C1A had a higher frequency of calcinosis cutis (*p*<0.01). There were no significant differences between anti-NT5C1A positive patients and negative patients with antisynthetase syndrome or IMNM. Of note, anti-NT5C1A positive dermatomyositis or antisynthetase syndrome patients did not show a higher association with interstitial lung disease. Also, anti-NT5C1A status did not correlate with treatment response in patients seropositive for anti-HMGCR antibody or anti-SRP antibody (*p*=0.61, *p*=0.53). Interestingly, among patients with dermatomyositis, antisynthetase syndrome, and IMNM that had an associated malignancy, all were seronegative for anti-NT5C1A. Among the 103 non-inflammatory muscle disease, VCP-related multi-system proteinopathy showed higher rate of anti-NT5C1A seropositivity compared with that in other non-inflammatory muscle diseases (*p*=0.03) (Table 1).

There were 48 patients with IBM whose anti-NT5C1A status had been retested at an interval of greater than one year. Anti-NT5C1A status did not change in 43/48 (90%) of patients. Among 22 patients who were negative for anti-NT5C1A, four patients (18%) converted to positive. There was no significant difference between these four patients and those whose antibody status remained negative regarding age at onset (*p*=0.6), time of onset to biopsy (*p*=0.63), or serum CK level (*p*=0.33).

## DISCUSSION

In this study, we explored the clinical utility of anti-NT5C1A antibody testing in a large number of patients with IIMs. Among patients with muscle diseases, the seropositivity of anti-NT5C1A was 63% sensitive and 85% specific for the diagnosis of IBM. This result compares with the results of previous studies, which suggested the range of sensitivity was 33% to 80% and that of specificity was 87% to 100% [2, 3, 5, 6, 7, 8, 21]. The Washington University Neuromuscular Clinical Laboratory clinically performs anti-NT5C1A antibody testing via western blot. The sensitivity of anti-NT5C1A for IBM with western blot was 61% in a previous study and 63% in this study [7]. Among studies using more than 10 samples from patients with IBM, the sensitivities of western blotting were consistently higher than that of ELISA assay unless a combination of ELISA assays were used to detect IgM, IgA, and IgG anti-NT5C1A antibodies [21].

Whether NT5C1A antibody positivity can be used to clinically support a diagnosis of IBM has been debated. We demonstrate in this study that ROC curve analysis using a linear regression model that included anti-NT5C1A status improved diagnostic accuracy. This analysis intentionally did not include biopsy data which may not be performed on some patients (Fig. 2 Curve 2). When ROC curve analysis also included the minimal biopsy feature of endomysial inflammation the sensitivity and specificity further improved (Fig. 2 Curve 3). These data support that the diagnosis of IBM does not rely on a single piece of clinical data and that anti-NT5C1A is an additional diagnostic tool. This is further highlighted by the fact that our data finds that when an IBM patient has finger flexion weakness and/or knee extension weakness, the presence of vacuoles is detected in 158 patients (65%) and focal invasion is detected in 128 patients (68%). Thus, the sensitivity of anti-NT5C1A alone (63%) is comparable to biopsy findings known to be characteristic for IBM, albeit the specificity of a biopsy with rimmed vacuoles is likely superior to anti-NT5C1A antibody positivity.

A previous report found that anti-NT5C1A antibody positive patients had a lower frequency of rimmed vacuoles [7]. Whether this is due to clinicians more readily ordering anti-NT5C1A antibody testing on patients without biopsy features regarded as specific for IBM (i.e. rimmed vacuoles) is not clear. This is consistent with our finding that 37 patients in our cohort with a clinical diagnosis of IBM had anti-NT5C1A testing without muscle biopsy and suggests that in clinical practice, clinicians are foregoing biopsy for less invasive testing such as serum autoantibodies.

Interestingly, the seropositivity for anti-NT5C1A in VCP-related multi-system proteinopathy was higher than that in other non-inflammatory muscle diseases. Some patients with clinico-pathologically defined IBM have been found to have pathogenic VCP mutations [22, 23]. Whether this finding is related to the presence of rimmed vacuoles seen in VCP-related multi-system proteinopathy patients remains to be established.

In IBM patients, anti-NT5C1A seropositivity has been previously associated with an increase in biopsies containing COX negative fibers [12]. Our study similarly found that anti-NT5C1A positive IBM patients had biopsies with an increased percentage of COX negative fibers. Notably anti-NT5C1A positivity also strongly correlated with the presence of endomysial inflammation even in non-IBM associated IIMs.

Previously, seropositivity for anti-NT5C1A has been reported to correlate with a more severe phenotype in IBM, dysphagia and facial weakness [11, 12]. Other reports have failed to find a difference between anti-NT5C1A positive and negative patients [24]. Although these three studies did not show the relevance between seropositivity and the distal upper limb/grip weakness, in our study, seropositive IBM patients showed more frequent involvement of finger flexion weakness. The different results may be derived from the difference of method to detect the antibody, inclusion criteria, or the number of patients evaluated.

This is the first assessment of anti-NT5C1A status correlated with the clinicopathologic features of antisynthetase syndrome and IMNM. Although there were patients who were seropositive for anti-NT5C1A, their clinicopathologic features were not different from those in the seronegative patients. Specifically pertaining to malignancy (a feature important for patient management and prognosis), seropositive patients showed no clear association in dermatomyositis, antisynthetase syndrome, and IMNM.

In this study, several IBM patients had repeat anti-NT5C1A testing. When a patient was negative for anti-NT5C1A, 18% seroconverted to positive in the repeat test for anti-NT5C1A, while 96% of patients who were seropositive for the antibody remained positive in the follow up test. In clinical practice, this information will be helpful when considering if repeat testing may be helpful in the situation when the first test was negative for anti-NT5C1A.

This study has limitations. First, this is a retrospective study of clinical records and some features which were not described in the clinical charts may not have been adequately captured. Second, more numbers of patients with dermatomyositis, antisynthetase syndrome, and IMNM are needed in order to discuss a more precise statistical significance. Nevertheless, this study demonstrates the clinical utility of anti-NT5C1A in idiopathic inflammatory myopathies. Specifically, anti-NT5C1A testing increases the diagnostic sensitivity and specificity for IBM when combined with limited clinical information. We suggest that anti-NT5C1A testing be included as part of future diagnostic algorithms distinguishing IBM from other IIMs.

## Author Contributions

C.I., A.R.F., N.A.G., R.C.B., T.M., A.P., and C.C.W. contributed to the conception and design of the study. C.I., A.R.F., N.A.G., S.R., J.C., Y.H., L.H.W., J.C.K., B.A.B., T.M., and C.C.W. contributed to data acquisition. C.I., S.R., M.W., and C.C.W. contributed to analysis of data. C.I. and C.C.W. contributed to drafting the manuscript. C.I., A.R.F., N.A.G., R.C.B., T.M., A.P., and C.C.W. contributed to revising the manuscript for intellectual content.

## Potential conflicts of interest

Nothing to report.

## Notes

**Study funding:** This research was supported by CureIBM, Catalyze a Cure and NIH grants K24AR073317 to CCW and K08AR075894 to ARF.

### Competing Interest Statement

The authors have declared no competing interest.

### Funding Statement

This research was supported by CureIBM, Catalyze a Cure and NIH grants K24AR073317 to CCW and K08AR075894 to ARF.

## REFERENCES

1. Weihl CC, Mammen AL. Sporadic inclusion body myositis - a myodegenerative disease or an inflammatory myopathy. Neuropathol Appl Neurobiol. 2017;43(1):82–91.

2. Pluk H, van Hoeve BJ, van Dooren SH, et al. Autoantibodies to cytosolic 5’-nucleotidase 1A in inclusion body myositis. Ann Neurol. 2013;73(3):397–407.

3. Larman HB, Salajegheh M, Nazareno R, et al. Cytosolic 5’-nucleotidase 1A autoimmunity in sporadic inclusion body myositis. Ann Neurol. 2013;73(3):408–418.

4. Amlani A, Choi MY, Tarnopolsky M, et al. Anti-NT5c1A Autoantibodies as Biomarkers in Inclusion Body Myositis. Front Immunol. 2019; 10:745.

5. Herbert MK, Stammen-Vogelzangs J, Verbeek MM, et al. Disease specificity of autoantibodies to cytosolic 5’-nucleotidase 1A in sporadic inclusion body myositis versus known autoimmune diseases. Ann Rheum Dis. 2016;75(4):696–701.

6. Kramp SL, Karayev D, Shen G, et al. Development and evaluation of a standardized ELISA for the determination of autoantibodies against NT5C1A (Mup44, NT5C1A) in sporadic inclusion body myositis. Auto Immun Highlights. 2016;7(1):16.

7. Lloyd TE, Christopher-Stine L, Pinal-Fernandez I, et al. Cytosolic 5’-Nucleotidase 1A As a Target of Circulating Autoantibodies in Autoimmune Diseases. Arthritis Care Res (Hoboken). 2016;68(1):66–71.

8. Muro Y, Nakanishi H, Katsuno M, Kono M, Akiyama M. Prevalence of anti-NT5C1A antibodies in Japanese patients with autoimmune rheumatic diseases in comparison with other patient cohorts. Clin Chim Acta. 2017; 472:1–4.

9. Rietveld A, van den Hoogen LL, Bizzaro N, et al. Autoantibodies to Cytosolic 5’-Nucleotidase 1A in Primary Sjögren’s Syndrome and Systemic Lupus Erythematosus. Front Immunol. 2018; 9:1200.

10. Yeker RM, Pinal-Fernandez I, Kishi T, et al. Anti-NT5C1A autoantibodies are associated with more severe disease in patients with juvenile myositis. Ann Rheum Dis. 2018;77(5):714–719.

11. Goyal NA, Cash TM, Alam U, et al. Seropositivity for NT5c1A antibody in sporadic inclusion body myositis predicts more severe motor, bulbar and respiratory involvement. J Neurol Neurosurg Psychiatry. 2016;87(4):373–378.

12. Lilleker JB, Rietveld A, Pye SR, et al. Cytosolic 5’-nucleotidase 1A autoantibody profile and clinical characteristics in inclusion body myositis. Ann Rheum Dis. 2017.

13. Rose MR, Group EIW. 188th ENMC International Workshop: Inclusion Body Myositis, 2-4 December 2011, Naarden, The Netherlands. Neuromuscul Disord. 2013;23(12):1044–1055.

14. Allenbach Y, Mammen AL, Benveniste O, Stenzel W, Group I-MNMW. 224th ENMC International Workshop:: Clinico-sero-pathological classification of immune-mediated necrotizing myopathies Zandvoort, The Netherlands, 14-16 October 2016. Neuromuscul Disord. 2018;28(1):87–99.

15. Hoogendijk JE, Amato AA, Lecky BR, et al. 119th ENMC international workshop: trial design in adult idiopathic inflammatory myopathies, with the exception of inclusion body myositis, 10-12 October 2003, Naarden, The Netherlands. Neuromuscul Disord. 2004;14(5):337–345.

16. Bucelli RC, Pestronk A. Immune myopathies with perimysial pathology: Clinical and laboratory features. Neurol Neuroimmunol Neuroinflamm. 2018;5(2): e434.

17. Connors GR, Christopher-Stine L, Oddis CV, Danoff SK. Interstitial lung disease associated with the idiopathic inflammatory myopathies: what progress has been made in the past 35 years? Chest. 2010;138(6):1464–1474.

18. Witt LJ, Curran JJ, Strek ME. The Diagnosis and Treatment of Antisynthetase Syndrome. Clin Pulm Med. 2016;23(5):218–226.

19. Alshehri A, Choksi R, Bucelli R, Pestronk A. Myopathy with anti-HMGCR antibodies: Perimysium and myofiber pathology. Neurol Neuroimmunol Neuroinflamm. 2015;2(4): e124.

20. Temiz P, Weihl CC, Pestronk A. Inflammatory myopathies with mitochondrial pathology and protein aggregates. J Neurol Sci. 2009;278(1-2):25–29.

21. Greenberg SA. Cytoplasmic 5’-nucleotidase autoantibodies in inclusion body myositis: Isotypes and diagnostic utility. Muscle Nerve. 2014;50(4):488–492.

22. Weihl CC, Baloh RH, Lee Y, et al. Targeted sequencing and identification of genetic variants in sporadic inclusion body myositis. Neuromuscul Disord. 2015;25(4):289–296.

23. Gang Q, Bettencourt C, Machado PM, et al. Rare variants in SQSTM1 and VCP genes and risk of sporadic inclusion body myositis. Neurobiol Aging. 2016; 47:218. e211-218.e219.

24. Felice KJ, Whitaker CH, Wu Q, et al. Sensitivity and clinical utility of the anti-cytosolic 5’-nucleotidase 1A (NT5C1A) antibody test in sporadic inclusion body myositis: Report of 40 patients from a single neuromuscular center. Neuromuscul Disord. 2018;28(8):660–664.

